# Low Coverage, Moderate Knowledge, And Limited Trust: An Assessment of Knowledge, Attitudes, And Perceptions Towards Indoor Residual Spraying in Masala, Ndola, Zambia

**DOI:** 10.64898/2026.09.04.26362228

**Authors:** Florence Mabaso, Hope Mwelwa, Ephraim Mapiki, Chileleko Mapiki

## Abstract

**Background:** Indoor Residual Spraying (IRS) is a proven vector control intervention for malaria prevention, yet community acceptance remains suboptimal in many settings. This study assessed the knowledge, attitudes, and perceptions towards IRS among residents of Masala Township in Ndola, Copperbelt Province, Zambia.

**Methods:** A community-based cross-sectional study was conducted among 282 systematically selected adult residents of Masala Township using a structured questionnaire administered through face-to-face interviews (response rate: 89.5%). Knowledge scores (range 0-6) were categorized as poor, moderate, or good. Attitudes were measured using Likert scale items (range 1-5) and categorized as positive, neutral, or negative. Data were analysed using descriptive statistics, chisquare tests, and binary logistic regression (p<0.05).

**Results:** IRS coverage in the last 12 months was low at 24.8% (70/282). Knowledge levels were moderate in 43.6%, with only 24.8% demonstrating good knowledge. Attitudes were neutral overall (mean score 3.2±0.8), with 29.8% positive, 43.6% neutral, and 26.6% negative. Perceptions of chemical safety were poor, with only 39.7% believing IRS chemicals are safe. Fear of side effects (46.2%) and limited of trust in the spraying process (31.6%) were the most commonly reported barriers to acceptance. Associated factors with positive attitudes included: past spraying experience (AOR=4.56, p<0.001), tertiary education (AOR=3.45, p=0.002), higher income (AOR=2.89, p=0.007), plastered walls (AOR=2.12, p=0.029), and better knowledge (AOR=1.42, p=0.003).

**Conclusion:** IRS coverage in Masala (24.8%) is substantially below the WHO-recommended 80% threshold. Fear of side effects and lack of trust were the main barriers to acceptance. Factors associated with positive attitudes included education, income, knowledge, wall type, past spraying experience, and trust in health workers. Given the cross-sectional design, these findings represent associations, not causal relationships. Urgent interventions addressing safety concerns through enhanced community sensitization and trust-building are needed to improve IRS uptake and progress towards Zambia’s malaria elimination goals.

## BACKGROUND

Malaria remains a leading public health challenge, particularly in sub-Saharan Africa, where the disease burden is disproportionately high. According to the World Health Organization (WHO), an estimated 249 million malaria cases and 608,000 deaths occurred globally in 2022, with over 95% of these cases reported in Africa (World Health Organization, 2023). While regions such as Southeast Asia and Latin America have reported significant declines in malaria transmission, sub-Saharan Africa remains the hardest hit (Bhatt et al., 2015).

Zambia, classified as a high-burden malaria country, continues to struggle with endemic transmission, particularly in regions such as the Copperbelt, where Masala is located (World Health Organization, 2023). According to the 2022 Zambia National Malaria Indicator Survey, malaria prevalence in Copperbelt Province is approximately 8.5%, with significant variation between urban and peri-urban areas. Ndola District, where Masala is located, reports some of the highest malaria case burdens in the province, accounting for over 20% of all malaria cases in Copperbelt (Zambia Ministry of Health, 2023). The National Malaria Elimination Strategic Plan (NMESP 2021-2026) aims to reduce malaria cases to below 5% prevalence by 2026, with Indoor Residual Spraying (IRS) being a key intervention (Zambia National Malaria Elimination Centre, 2021).

To mitigate malaria transmission, Zambia has implemented various malaria control strategies, including Indoor Residual Spraying (IRS), insecticide-treated nets (ITNs), and case management with artemisinin-based combination therapy (ACTs) (Zambia National Malaria Elimination Centre, 2021). IRS involves the application of long-acting residual insecticides to the interior walls and ceilings of houses. Mosquitoes that rest on these treated surfaces after feeding are killed, reducing both vector density and malaria transmission. The insecticide remains effective for 3-6 months, depending on the product used and wall surface type (World Health Organization, 2020). Commonly used insecticides include pyrethroids and organophosphates such as Actellic 300CS. A Cochrane systematic review estimated that IRS can reduce malaria incidence by up to 62% when coverage exceeds 80% of targeted households and when insecticides remain effective against local vector populations (Pluess et al., 2010).

Despite its effectiveness, factors such as community acceptability, knowledge levels, and perceptions influence the success of IRS programs (Suuron et al., 2020). Acceptance of IRS varies due to factors such as misinformation, religious beliefs, concerns about chemical exposure, and household structural challenges (Suuron et al., 2020).

IRS has been implemented in Zambia since the early 2000s as part of the Integrated Vector Management (IVM) strategy (Chanda et al., 2008). The Zambian government, in collaboration with the World Health Organization, the U.S. President’s Malaria Initiative (PMI), and the Global Fund, has expanded IRS coverage to high-risk areas, including parts of Copperbelt Province (President’s Malaria Initiative, 2024). Key components of IRS implementation in Zambia include: Targeting high-burden areas with high malaria transmission, use of longlasting insecticides including organophosphates (e.g., Actellic 300CS) and pyrethroids (Chanda et al., 2013), community sensitization conducted before each spray cycle to increase uptake and routine monitoring and evaluation tracking insecticide resistance and effectiveness (Moss et al., 2012)

However, insecticide resistance poses an increasing challenge to IRS effectiveness in Zambia. Pyrethroid resistance is now widespread in Anopheles vector populations across the country, prompting a shift towards organophosphate insecticides such as Actellic 300CS in some areas (Moss et al., 2012).

Several factors influence the acceptability and uptake of IRS in many communities, including knowledge and awareness, cultural and religious beliefs, perceptions of safety and effectiveness, and socio-economic and structural barriers (Oladipo et al., 2022). Studies from other settings have documented that fear of side effects, mistrust of spraying chemicals, and concerns about stains and odors are associated with lower participation in vector control campaigns (Musoke et al., 2015; Buttenheim et al., 2013; Rodriguez et al., 2006).

The Copperbelt remains a hotspot for malaria in Zambia, and in response, the Government has designed a Copperbelt Malaria Elimination Program (Ministry of Health, 2021). This program includes malaria control activities such as IRS. However, a study conducted in Masala, Ndola revealed that only 24.8% of Masala residents had their houses sprayed in the past 12 months, compared to 68.7% of respondents in Nkwazi (Vivian, 2019). In addition, Masala reports an endemic prevalence of malaria cases with more under-5 children affected. Masala Clinic’s Health Management Information System (HMIS) data indicate that malaria accounts for approximately 35% of all outpatient visits, with children under five accounting for over 40% of confirmed malaria cases (Masala Clinic, 2025).

Despite the recognized effectiveness of IRS, coverage in Masala (24.8%) is substantially below the WHO-recommended 80% threshold (World Health Organization, 2021). However, the specific factors driving low coverage—whether knowledge gaps, negative attitudes, safety concerns, or structural barriers— remain poorly understood in this peri-urban context. Understanding these factors is essential for designing effective interventions to increase IRS acceptance and, ultimately, reduce malaria transmission. A KAP study can identify specific knowledge gaps, misconceptions, attitudinal barriers, and perceptual concerns that may be contributing to low IRS acceptance. These insights can inform the design of targeted health education messages, sensitization strategies, and programmatic adaptations to address community-specific barriers. This study therefore aimed to assess the knowledge, attitudes, and perceptions towards IRS among Masala residents to inform programmatic decision-making.

## METHODS

### Study Design and Setting

This community-based cross-sectional study was conducted in Masala Township, a densely populated urban area in Ndola District, Copperbelt Province, Zambia. Masala Township is predominantly from middle and low socioeconomic backgrounds. The area is characterized by high malaria prevalence and challenges such as overcrowding at health facilities and reliance on self-treatment practices for malaria. According to the official 2022 Census of Population and Housing released by the Zambia Statistics Agency (ZamStats), the population of Masala Ward in Ndola is 10,127 people.

### Study Population and Sampling

The target population consisted of adult residents of Masala Township, Ndola, Zambia, who were directly or indirectly affected by malaria and were potential beneficiaries or participants in IRS programs. Inclusion criteria were households where the head or representative provided informed consent to participate, households available at the time of data collection, and households located in Masala Township, Ndola, Copperbelt. Exclusion criteria were households that did not provide consent, households not available during the data collection period and non-residential structures.

Sample size was calculated using Cochran’s formula: n = (Z² × p × q) / e², where Z=1.96, p=0.248 (from Vivian 2019), q=1-p=0.752, and e=0.05. This yielded a required sample of 287 participants.

Of the 287 households targeted, 282 completed the survey (98.3% of target), yielding a final sample of 282. The shortfall was due to refusal to participate.

This study employed a systematic random sampling method to select 287 households in Masala. Using an average household size of 4 persons, the estimated total number of households was calculated as approximately 2,532 (10,127/4). A sampling interval (k) of 9 was derived by dividing the total households by the target sample size (2,532 / 287). A random starting point was selected between 1 and 9, and thereafter every 9th household was systematically selected until the study sample size was achieved.

### Data Collection Tools

Data were collected using a structured, interviewer-administered questionnaire adapted from similar KAP studies on malaria interventions (16,17). The questionnaire was pretested on 15 participants from Masala Township to assess clarity, comprehension, and cultural appropriateness. Based on pretesting, minor wording adjustments were made. Research assistants were trained over two days on study objectives, questionnaire administration, ethical considerations, and interview techniques. The questionnaire was administered in English and the local language (Bemba), with translation performed by trained bilingual research assistants using a forward-backward translation method to ensure semantic equivalence.

Data collection was conducted from April to August 2025, covering the period before and during the typical malaria transmission season in Zambia. Data collection was conducted using KoBoToolbox on electronic tablets, where enumerators administered the structured questionnaire through the KoBoCollect mobile app. This method ensured systematic coverage of the study area, minimized selection bias, and allowed for efficient real-time data capture and quality control.

The questionnaire captured: (1) demographic characteristics (age, gender, education, occupation, wall material, household income, household size, presence of children under 5, presence of pregnant women); (2) knowledge of IRS benefits and information sources; (3) attitudes towards IRS; and (4) perceptions towards safety and effectiveness of IRS.

A knowledge score was computed from responses to questions regarding awareness of IRS, belief in IRS effectiveness, and knowledge of IRS benefits. Scores ranged from 0 to 6, with higher scores indicating greater knowledge. Knowledge was assessed using 6 questions: (1) Heard about IRS, (2) IRS is effective, (3) IRS reduces mosquito populations, (4) IRS lowers malaria transmission, (5) IRS provides long-term protection, and (6) IRS improves community health. Each correct response scored 1 point (maximum 6). For analysis, scores were categorized as poor knowledge (0-2 correct), moderate knowledge (3-4 correct), and good knowledge (5-6 correct). Attitude was measured using a 5-point Likert scale ranging from strongly agree (5) to strongly disagree (1). Composite attitude scores were calculated and categorized as positive (mean ≥ 3.5), neutral (mean 2.5–3.4), or negative (mean ≤ 2.4).

### Validity and Reliability

To ensure validity, a preliminary review of the questionnaire was conducted with a small group of non-experts (community members) to assess clarity, readability, and relevance. In addition, a panel of experts (malaria control specialists, epidemiologists, and public health practitioners) evaluated whether the questionnaire adequately covered all dimensions of IRS-related knowledge, attitudes, and perceptions. Adjustments were made to eliminate redundant or irrelevant items.

To ensure reliability, Cronbach’s alpha was calculated for multi-item scales to ensure homogeneity. A value of □ ≥ 0.7 confirmed that items measuring the same construct were interrelated. The pretesting with 15 participants from Masala Township identified ambiguities, cultural misinterpretations, or technical flaws, and feedback was used to refine question phrasing and structure.

### Variables and Definitions

The primary outcome variables were knowledge level, attitude category, and perception regarding IRS. Behavioral variables included ever allowed spraying in the last 12 months, confidence in IRS, and belief that IRS should continue.

Definitions used in this study included: Indoor Residual Spraying (IRS): A malaria prevention method involving the application of long-lasting insecticides to interior walls and surfaces of houses where mosquitoes are likely to rest; Residual Insecticide: A long-lasting chemical that remains effective in killing insects for several months after application; Coverage: The proportion of households that reported ever allowing their houses to be sprayed under an IRS program, with effective programs typically aiming for over 80% coverage; and Insecticide Resistance: The ability of mosquitoes to survive exposure to a particular insecticide, which can reduce the effectiveness of IRS.

### Statistical Analysis

Data were entered into KoBoToolbox and exported to Microsoft Excel for cleaning, then analysed using IBM SPSS Version 24.0. Descriptive statistics (frequencies, percentages, means, standard deviations) were computed for all variables. Knowledge of IRS was assessed using six questions adapted from previous KAP studies on malaria interventions (Aongola et al., 2022; Jumbam et al., 2020). The questions assessed: (1) awareness of IRS, (2) belief in IRS effectiveness, (3) knowledge that IRS reduces mosquito populations, (4) knowledge that IRS lowers malaria transmission, (5) knowledge that IRS provides long-term protection, and (6) knowledge that IRS improves community health. Each correct response was scored 1 point, with “Not sure” responses scored as incorrect. The total knowledge score ranged from 0 to 6, with higher scores indicating greater knowledge. For analysis, scores were categorized as poor knowledge (0-2 correct), moderate knowledge (3-4 correct), and good knowledge (5-6 correct). The categorization was based on the distribution of scores and previous studies using similar scoring methods (Ngoma et al., 2026; Musoke et al., 2015). Cronbach’s alpha for the six-item knowledge scale was 0.72, indicating acceptable internal consistency.

Likert-scale attitude items were assessed for reliability using Cronbach’s alpha (□ ≥ 0.70), with composite mean scores calculated and categorized into positive, neutral, or negative attitudes. Perceptions regarding safety and effectiveness were analyzed using frequencies and percentages. Trust in health workers was measured using a single Likert scale item: ‘I trust health workers who conduct IRS’ (Table 6, item c). This was treated as a continuous variable (1=strongly disagree to 5=strongly agree) in the regression model.

Bivariate associations between categorical variables were assessed using chisquare tests of independence. Variables with p<0.20 in bivariate analysis were entered into a binary logistic regression model using a forward stepwise (likelihood ratio) method to identify factors independently associated with positive attitudes towards IRS. The model showed good fit (Hosmer-Lemeshow □^2^=6.34, p=0.609) and explained approximately 34.2% of the variance in attitudes. Adjusted odds ratios (AOR) with 95% confidence intervals (CI) were reported. Statistical significance was set at p<0.05.

There were no missing data for key outcome variables; missing demographic data (<5%) were handled using listwise deletion.

### Conceptual framework Diagram

**Fig 1:**
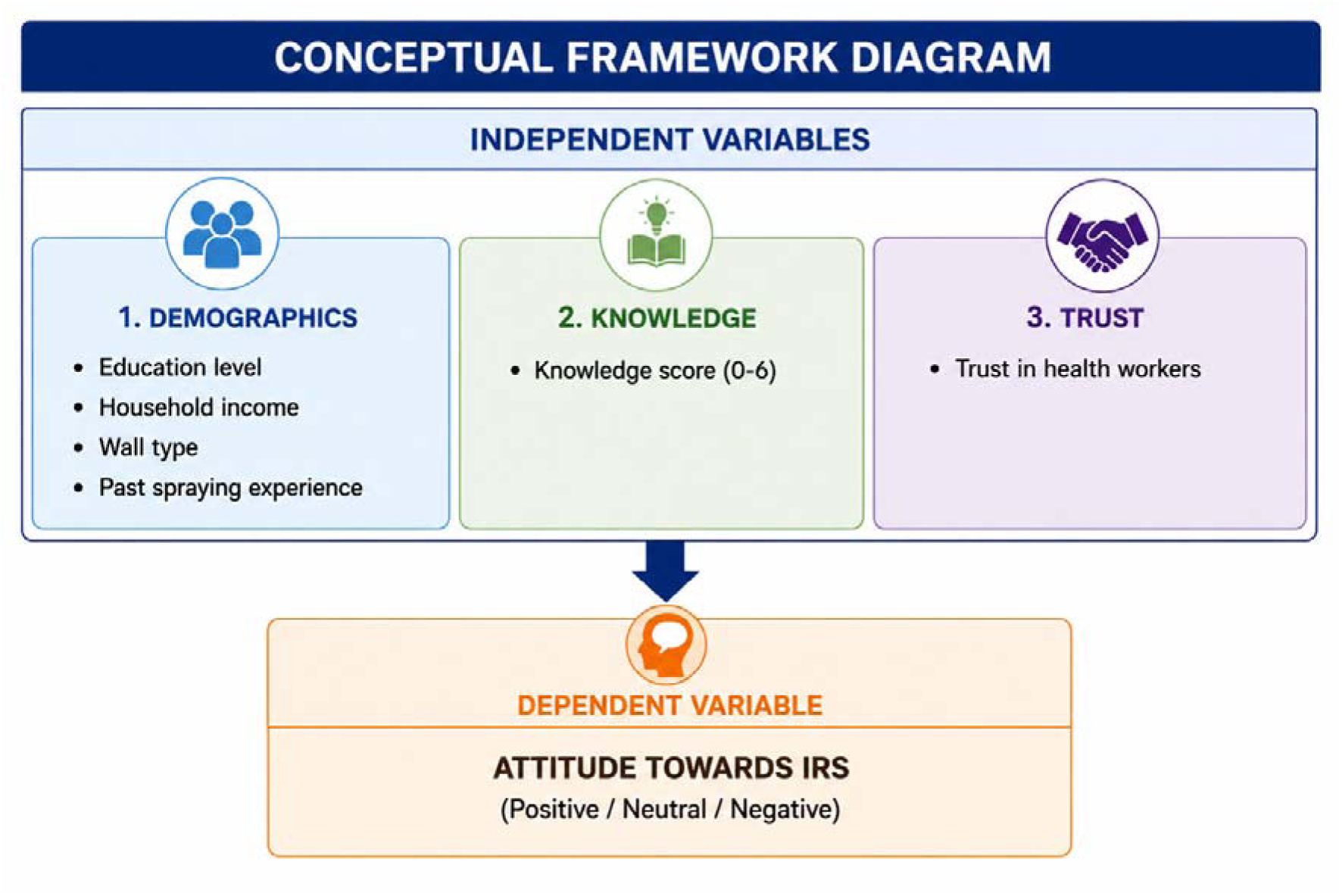
Conceptual framework Diagram -Attitudes towards IRS.png

### Ethical Considerations

Ethical approval was obtained from the Chreso University Research Ethics Committee (CUREC) (ref no. 22379-05-2025) prior to commencing the study. Permission was also obtained from local community authorities. All participants provided written informed consent. Participants were fully informed of the study’s purpose, procedures, and their right to withdraw at any time without consequences. Privacy was maintained during questionnaire administration. Data were anonymized and stored securely, with access restricted to the research team. Confidentiality was ensured throughout the study. Community engagement was prioritized throughout the study to respect local values, culture, and traditions, fostering trust and collaboration.

## RESULTS

### Response Rate

A total of 315 households were approached for participation in the study. Of these, 282 households provided complete responses, yielding a response rate of 89.5%. This represents 98.3% of the target sample of 287.

### Demographic Characteristics

The demographic characteristics of the 282 respondents are summarised in Table 1. The majority of respondents were aged 26-35 years (31.6%), females constituted the majority at 56.0%, regarding education, 44.0% had secondary education, 27.0% had primary education, 18.1% had tertiary education, and 11.0% had no formal education.

**Table 1.**
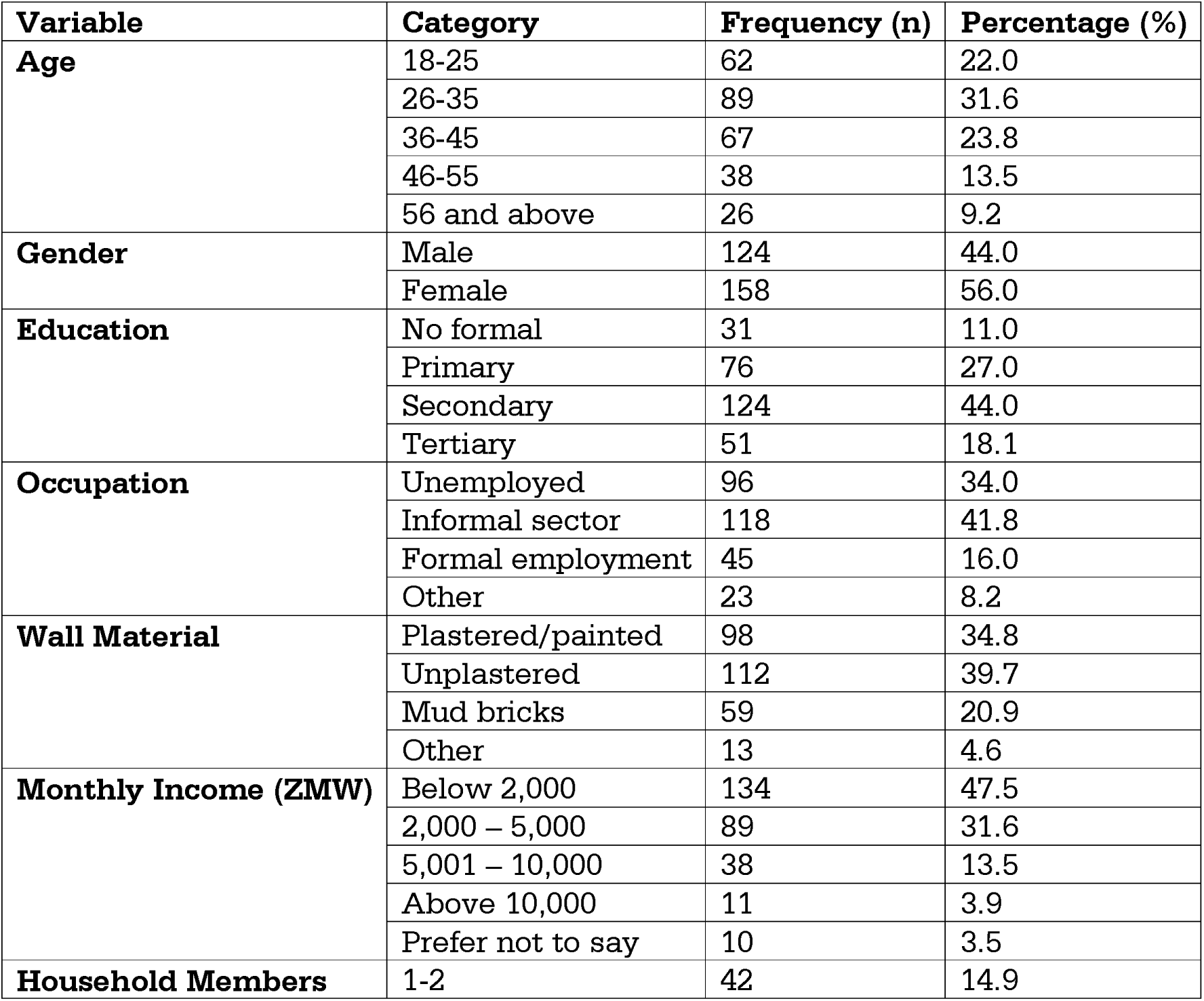
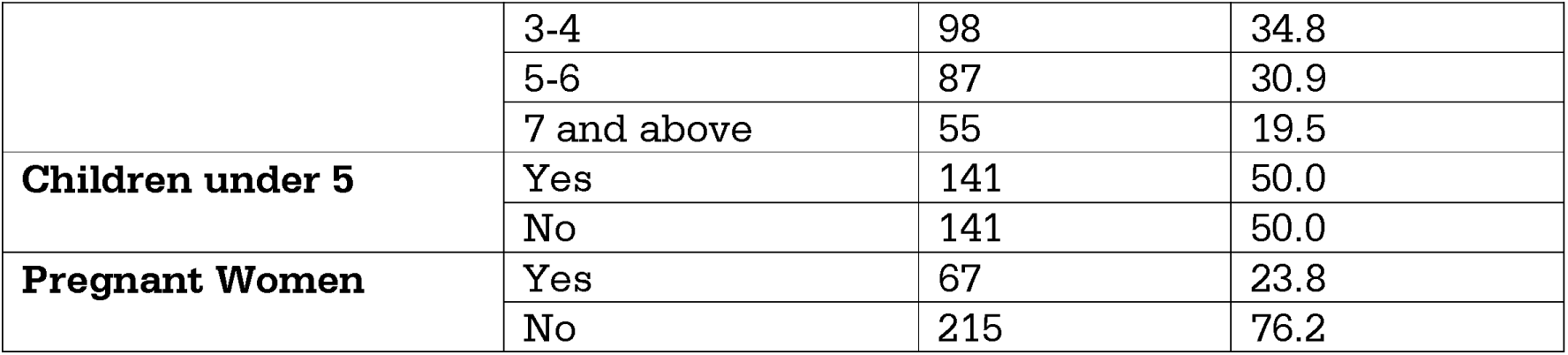
Demographic Characteristics of Respondents (n=282)

| **Variable** | **Category** | **Frequency (n)** | **Percentage (%)** |
| --- | --- | --- | --- |
| **Age** | 18-25 | 62 | 22.0 |
|  | 26-35 | 89 | 31.6 |
|  | 36-45 | 67 | 23.8 |
|  | 46-55 | 38 | 13.5 |
|  | 56 and above | 26 | 9.2 |
| **Gender** | Male | 124 | 44.0 |
|  | Female | 158 | 56.0 |
| **Education** | No formal | 31 | 11.0 |
|  | Primary | 76 | 27.0 |
|  | Secondary | 124 | 44.0 |
|  | Tertiary | 51 | 18.1 |
| **Occupation** | Unemployed | 96 | 34.0 |
|  | Informal sector | 118 | 41.8 |
|  | Formal employment | 45 | 16.0 |
|  | Other | 23 | 8.2 |
| **Wall Material** | Plastered/painted | 98 | 34.8 |
|  | Unplastered | 112 | 39.7 |
|  | Mud bricks | 59 | 20.9 |
|  | Other | 13 | 4.6 |
| **Monthly Income (ZMW)** | Below 2,000 | 134 | 47.5 |
|  | 2,000 – 5,000 | 89 | 31.6 |
|  | 5,001 – 10,000 | 38 | 13.5 |
|  | Above 10,000 | 11 | 3.9 |
|  | Prefer not to say | 10 | 3.5 |
| **Household Members** | 1-2 | 42 | 14.9 |
|  | 3-4 | 98 | 34.8 |
|  | 5-6 | 87 | 30.9 |
|  | 7 and above | 55 | 19.5 |
| **Children under 5** | Yes | 141 | 50.0 |
|  | No | 141 | 50.0 |
| **Pregnant Women** | Yes | 67 | 23.8 |
|  | No | 215 | 76.2 |

In terms of occupation, 41.8% of respondents were engaged in the informal sector, while 34.0% were unemployed. The majority of households (39.7%) had unplastered walls, while 34.8% had plastered/painted walls and 20.9% had mud brick walls. Household income was generally low, with 47.5% earning below ZMW 2,000 per month. Most households had 3-4 members (34.8%) or 5-6 members (30.9%). Half of the households (50.0%) had children under five years, and 23.8% had pregnant women residing in the household.

### Knowledge of IRS

The majority of respondents (70.2%) had heard about IRS, with health workers (39.7%) and radio/television (31.6%) being the primary information sources. While over half (55.3%) believed IRS is effective, knowledge of specific benefits varied considerably: 51.4% knew IRS reduces mosquito populations, 47.5% knew it lowers malaria transmission, but only 31.6% recognized long-term protection and just 23.8% knew it improves community health. Overall, the mean knowledge score was 3.2 (±1.6) out of 6, with the majority having moderate knowledge (43.6%), while only 24.8% demonstrated good knowledge and 31.6% had poor knowledge. This indicates that while general awareness is moderate, in-depth understanding of IRS benefits remains limited, particularly regarding broader community health impacts.

**Table 2.**
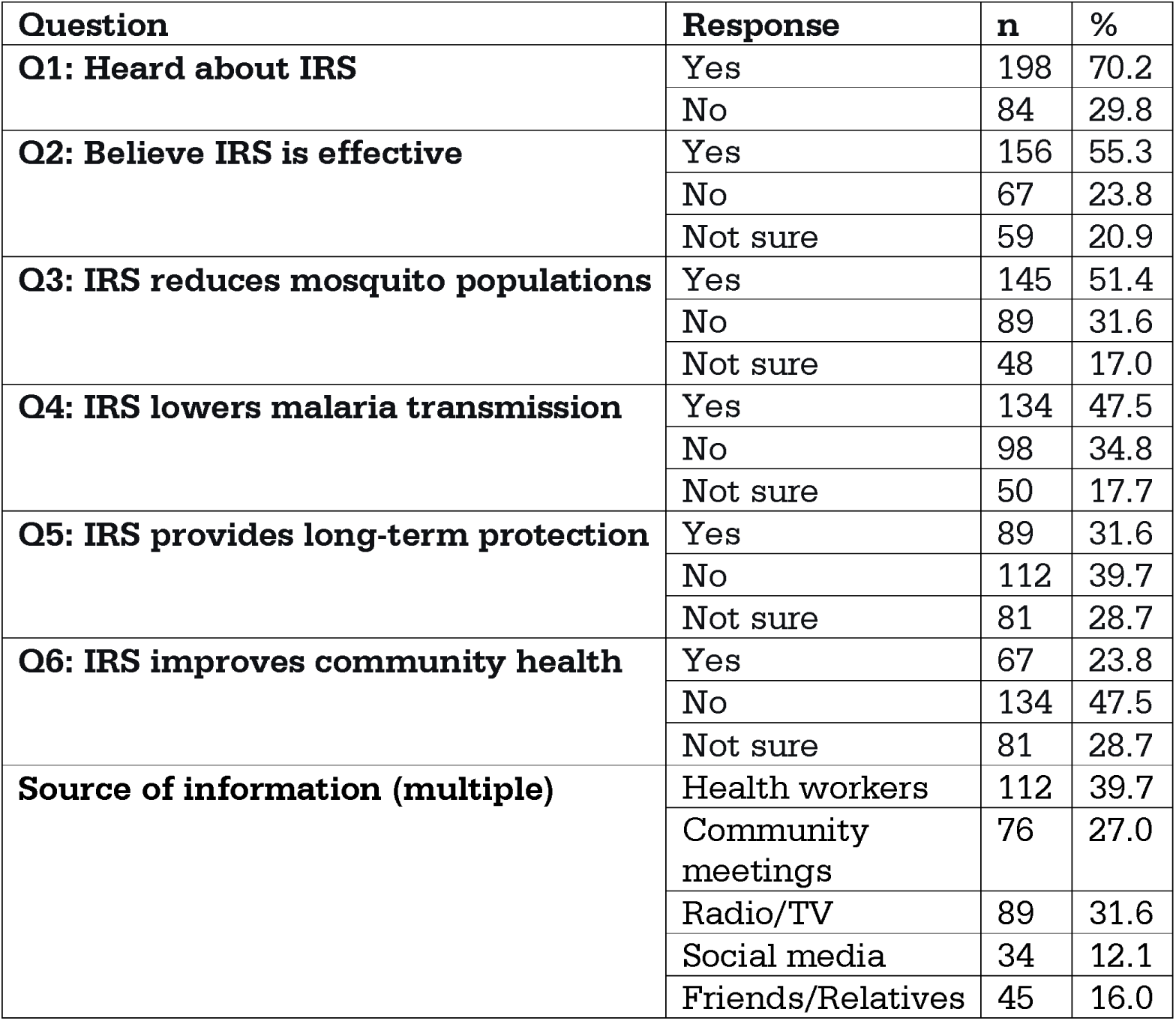
Knowledge of Respondents on IRS (n=282)

Overall knowledge scores revealed that the majority of respondents had moderate knowledge (43.6%), while 31.6% had poor knowledge and only 24.8% had good knowledge. The mean knowledge score was 3.2 (±1.6) out of a possible 6, indicating moderate levels of knowledge about IRS among Masala residents. Table 3 presents the overall knowledge levels.

**Table 3.** Overall Knowledge Levels on IRS (n=282)

| Knowledge Level | Score Range | n | % |
| --- | --- | --- | --- |
| Poor | 0-2 correct | 89 | 31.6 |
| Moderate | 3-4 correct | 123 | 43.6 |
| Good | 5-6 correct | 70 | 24.8 |
| **Mean Knowledge Score (SD)** | **3.2 ($\pm 1.6$)** |  |  |

**Fig 2:**
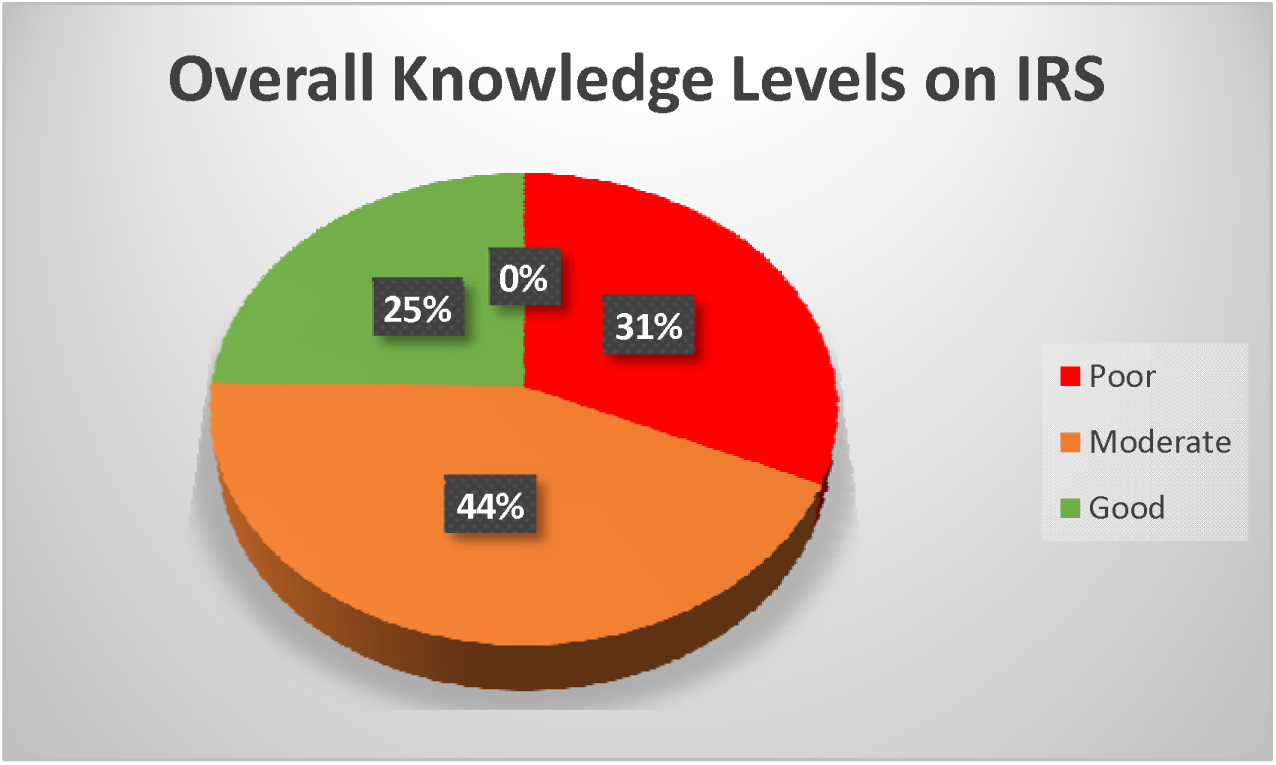
Overall Knowledge Levels on IRS

### Attitudes Towards IRS

The attitudes of respondents towards IRS are presented in Tables 4 and 5. Only 24.8% of respondents reported ever allowing their houses to be sprayed under an IRS programme in the last 12 months, while 75.2% had never participated. Among those who refused spraying (n=212), the most common reasons were fear of side effects (46.2%) and lack of trust in the spraying process (31.6%), followed by lack of awareness (26.4%) and religious/cultural beliefs (16.0%).

**Table 4.** Attitudes of Respondents Towards IRS (n=282)

| Question | Response | n | % |
| --- | --- | --- | --- |
| **Ever allowed spraying in the last 12 months** | Yes | 70 | 24.8 |
|  | No | 212 | 75.2 |
| **Reason for refusal (multiple, n=212)** | Fear of side effects | 98 | 46.2 |
|  | Religious/cultural beliefs | 34 | 16.0 |
|  | Lack of trust in process | 67 | 31.6 |
|  | Lack of awareness | 56 | 26.4 |
|  | Other | 23 | 10.8 |
| **Confidence in IRS** | Very confident | 45 | 16.0 |
|  | Somewhat confident | 89 | 31.6 |
|  | Neutral | 56 | 19.8 |
|  | Not confident | 56 | 19.8 |
|  | No confidence at all | 36 | 12.8 |
| **Should IRS continue** | Yes | 167 | 59.2 |
|  | No | 56 | 19.9 |
|  | Not sure | 59 | 20.9 |

**Table 5.** Likert Scale Attitude Items Towards IRS (n=282)

| Statement | Mean (SD) | Interpretation |
| --- | --- | --- |
| a) IRS is safe for my family | 3.1 ($\pm 1.2$) | Neutral |
| b) IRS is effective in reducing malaria | 3.4 ($\pm 1.1$) | Neutral/Agree |
| c) I trust health workers who conduct IRS | 3.2 ($\pm 1.3$) | Neutral |
| d) IRS causes unpleasant odors | 3.8 ($\pm 1.0$) | Agree |
| e) IRS leaves stains on walls | 3.6 ($\pm 1.1$) | Agree |
| f) I would allow IRS again | 3.0 ($\pm 1.4$) | Neutral |
| g) IRS is a waste of resources | 3.3 ($\pm 1.2$) | Neutral |
| h) I feel protected after IRS | 3.5 ($\pm 1.1$) | Agree |
| **Composite Attitude Score (Mean)** | **3.2 ($\pm 0.8$)** | **Neutral** |

Confidence in IRS as a malaria prevention method was mixed: 31.6% were somewhat confident, while 19.8% were neutral, 19.8% were not confident, and 12.8% had no confidence at all. Only 16.0% expressed being very confident. Despite this, the majority (59.2%) believed IRS should be continued in their community.

Of particular note, the item ‘I would allow IRS again’ had a mean score of 3.0 (SD=1.4), indicating that respondents were equally split on whether they would participate in future spraying campaigns.

**Fig 3:**
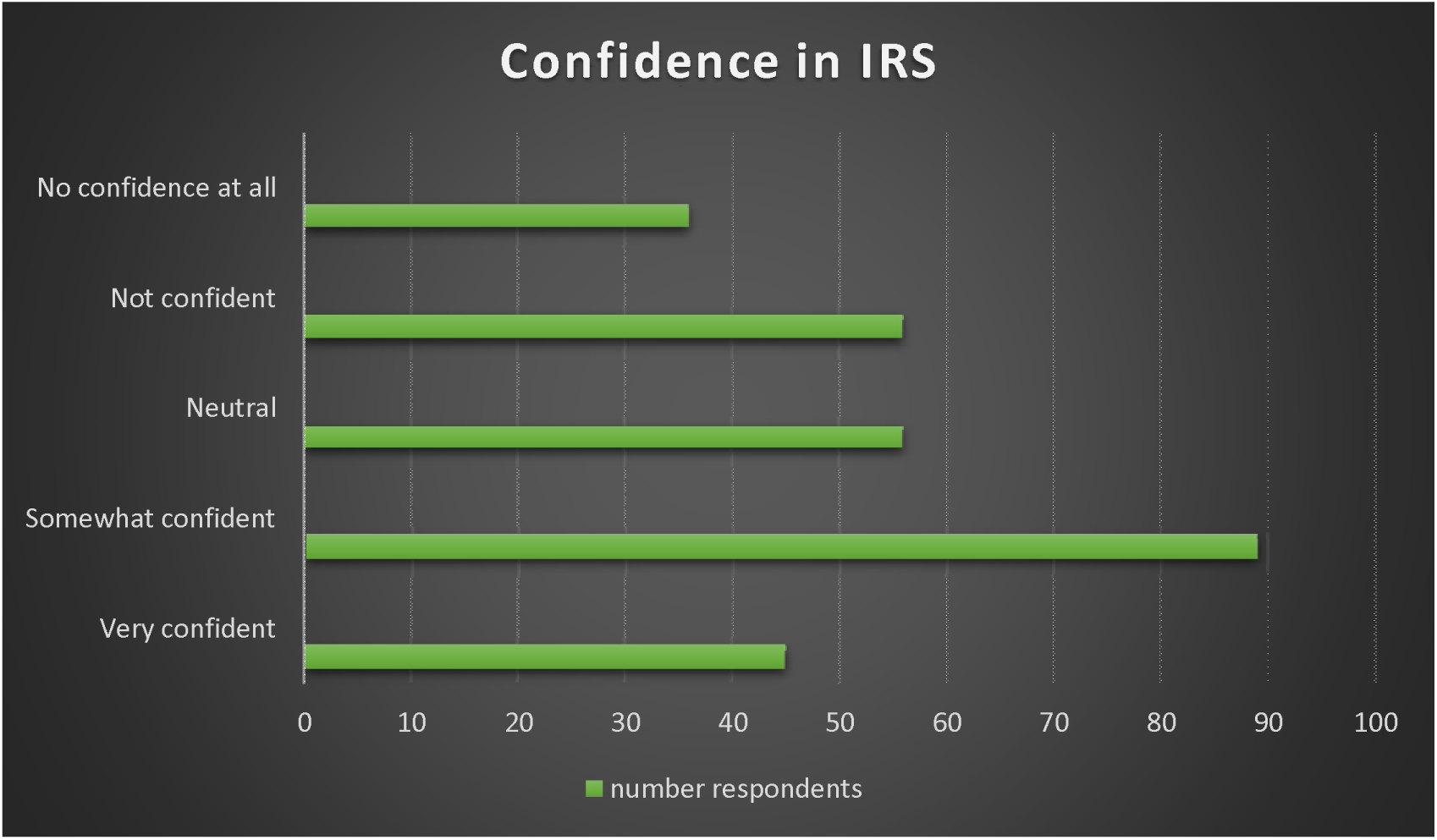
Confidence in IRS

The Likert scale attitude items (Table 5) revealed that respondents generally agreed that IRS causes unpleasant odors (mean=3.8) and leaves stains on walls (mean=3.6). They also agreed that they feel protected after IRS (mean=3.5). However, attitudes towards IRS safety (mean=3.1), effectiveness (mean=3.4), and trust in health workers (mean=3.2) were neutral. The composite mean attitude score was 3.2 (±0.8), indicating a neutral overall attitude towards IRS.

Categorisation of attitudes (Table 6) showed that 43.6% of respondents had neutral attitudes, while 29.8% had positive attitudes and 26.6% had negative attitudes towards IRS.

**Table 6.** Attitude Categorization (n=282)

| Attitude Category | n | % |
| --- | --- | --- |
| Positive (mean $\geq 3.5$) | 84 | 29.8 |
| Neutral (mean 2.5 – 3.4) | 123 | 43.6 |
| Negative (mean $\leq 2.4$) | 75 | 26.6 |

**Fig 4:**
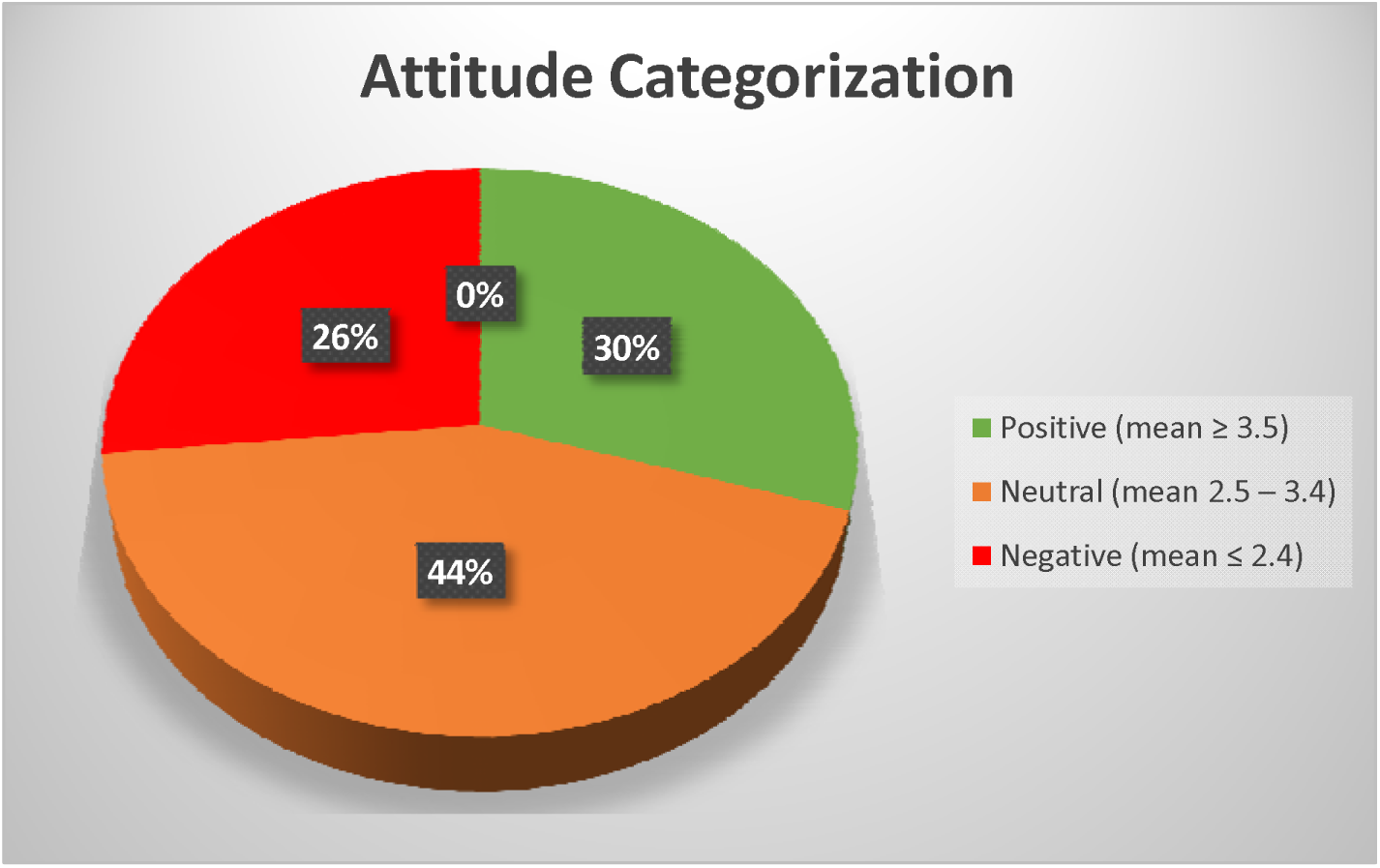
Attitude Categorization

### Perceptions Towards Safety and Effectiveness of IRS

Perceptions regarding the safety and effectiveness of IRS are shown in Table 7. Only 39.7% of respondents believed that the chemicals used in IRS are safe for humans, while 34.8% believed they were not safe and 25.5% were unsure. The most common concerns expressed were health risks (47.5%), environmental impact (31.6%), and doubts about effectiveness (27.0%). Notably, 19.9% of respondents reported having no concerns about IRS. The majority of respondents (60.3%) either believed that IRS chemicals are not safe for humans or were unsure about their safety, indicating widespread safety concerns.

**Table 7.**
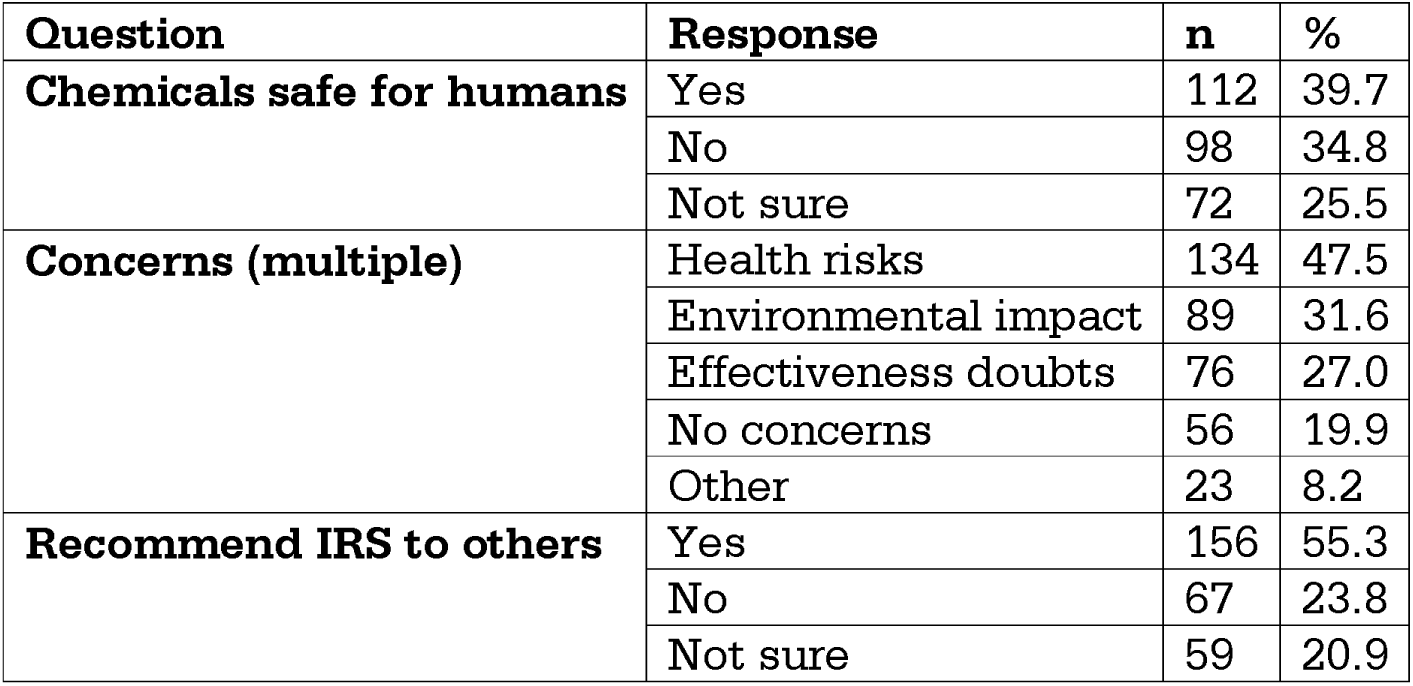
Perceptions of Respondents Towards Safety and Effectiveness of IRS (n=282)

Interestingly, 55.3% would recommend IRS to others, despite only 39.7% believing the chemicals are safe. This suggests that many respondents may see value in IRS despite lingering safety concerns.

**Fig 5:**
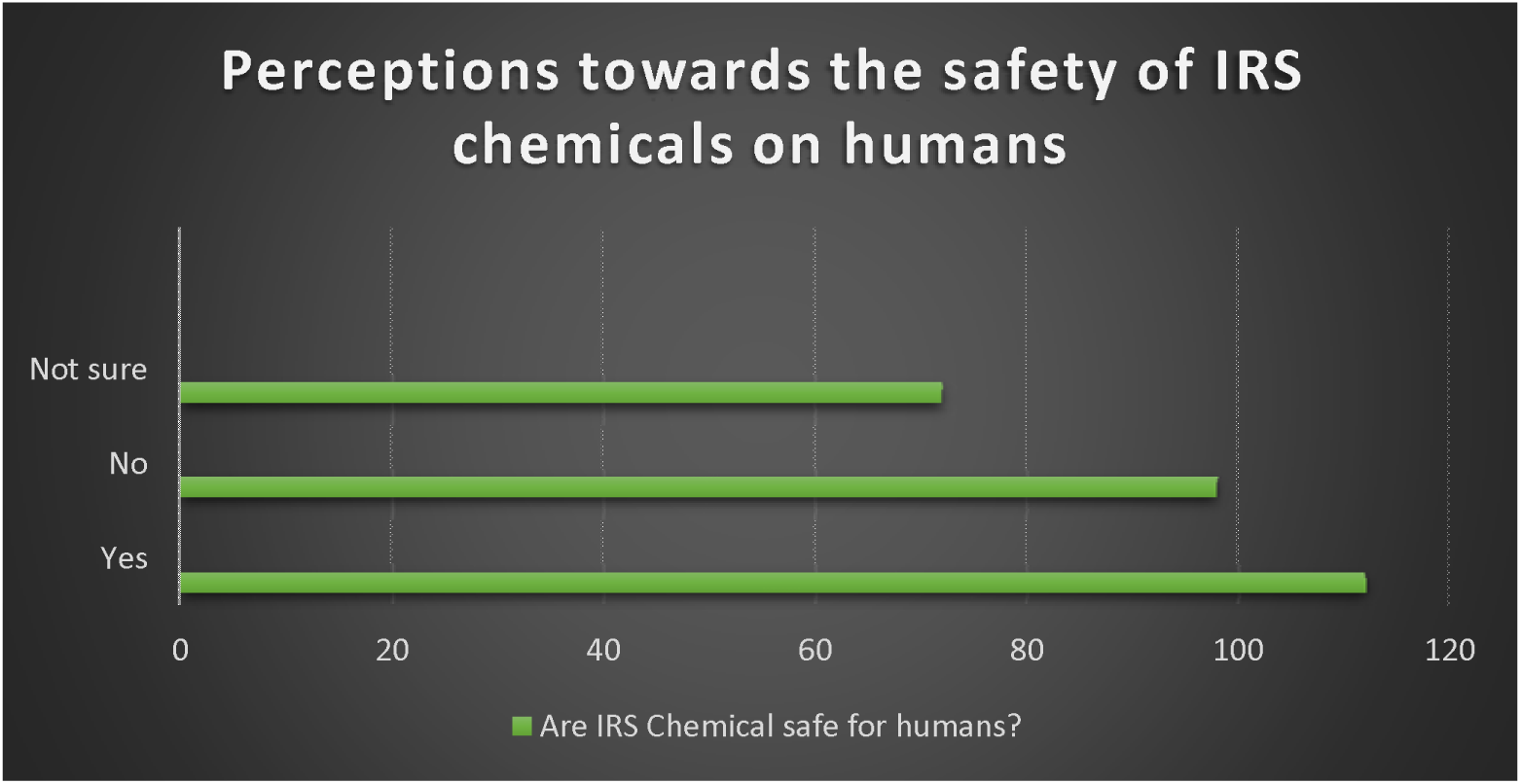
Perceptions towards the safety of IRS chemicals on humans

### Bivariate Associations

Table 8 presents bivariate associations between demographic factors and knowledge/attitudes using chi-square tests. Knowledge was significantly associated with age [□^2^(df=4)=15.67, p=0.042], education [□^2^(df=3)=22.34, p=0.011], occupation [□^2^(df=3)=18.92, p=0.045], and income [□^2^(df=3)=19.45, p=0.034]. Attitudes were significantly associated with age [□^2^(df=4)=16.23, p=0.038], education [□^2^(df=3)=20.89, p=0.023], wall material [□^2^(df=3)=16.78, p=0.031], and income [□^2^(df=3)=21.34, p=0.029]. Gender, presence of children under five, and presence of pregnant women showed no significant associations (p>0.05). Occupation approached but did not reach statistical significance for attitudes [□^2^(df=3)=14.56, p=0.056]. Occupation was significantly associated with knowledge but not attitude (p=0.056), suggesting that knowledge and attitudes may be influenced by different factors.

**Table 8.** Association Between Demographic Factors and Knowledge/Attitudes.

| Variable | Knowledge Level | Attitude Category |
| --- | --- | --- |
| Age | $\chi^2(df=8)=15.67$, **$p=0.042^*$** | $\chi^2(df=8)=16.23$, **$p=0.038^*$** |
| Gender | $\chi^2(df=2)=2.34$, $p=0.321$ | $\chi^2(df=2)=1.89$, $p=0.412$ |
| Education | $\chi^2(df=6)=22.34$, **$p=0.011^*$** | $\chi^2(df=6)=20.89$, **$p=0.023^*$** |
| Occupation | $\chi^2(df=6)=18.92$, **$p=0.045^*$** | $\chi^2(df=6)=14.56$, $p=0.056$ |
| Wall Material | $\chi^2(df=6)=10.45$, $p=0.078$ | $\chi^2(df=6)=16.78$, **$p=0.031^*$** |
| Income | $\chi^2(df=6)=19.45$, **$p=0.034^*$** | $\chi^2(df=6)=21.34$, **$p=0.029^*$** |
| Children under 5 | $\chi^2(df=1)=1.23$, $p=0.412$ | $\chi^2(df=1)=4.56$, $p=0.089$ |
| Pregnant women | $\chi^2(df=1)=0.89$, $p=0.567$ | $\chi^2(df=1)=2.34$, $p=0.234$ |

\*Significant at $p < 0.05$

### Factors associated with Positive Attitude Towards IRS

Binary logistic regression was performed to identify independent predictors of positive attitude towards IRS (Table 9). The model was adequate (HosmerLemeshow □^2^=6.34, p=0.609) and explained 34.2% of the variance in attitudes (Nagelkerke R²=0.342). The overall model correctly predicted 78.4% of cases.

**Table 9.** Binary Logistic Regression Predicting Positive Attitude Towards IRS (n=282)

| Predictor | Reference Category | Crude OR (95% CI) | p-value | Adjusted OR (95% CI) | p-value |
| --- | --- | --- | --- | --- | --- |
| **Education** | No formal education |  |  |  |  |
| Tertiary |  | 3.12 (1.45–6.71) | 0.004 | 3.45 (1.56–7.63) | **0.002\*** |
| **Income** | Below ZMW 2,000 |  |  |  |  |
| Above ZMW 5,000 |  | 2.45 (1.18–5.09) | 0.016 | 2.89 (1.34–6.21) | **0.007\*** |
| **Knowledge Score** | Per 1-point increase | 1.38 (1.10–1.73) | 0.005 | 1.42 (1.12–1.79) | **0.003\*** |
| **Wall Material** | Mud brick walls |  |  |  |  |
| Plastered/Painted |  | 1.98 (1.02–3.84) | 0.043 | 2.12 (1.08–4.17) | **0.029\*** |
| **Past Spraying** | No |  |  |  |  |
| Yes |  | 4.12 (1.98–8.57) | <0.001 | 4.56 (2.12–9.81) | **<0.001\*** |
| **Trust in Health Workers** | Per 1-point increase | 1.34 (1.02–1.76) | 0.034 | 1.38 (1.05–1.82) | **0.021\*** |

*Significant at p < 0.05*

The following factors significantly predicted positive attitudes:

- Tertiary education (AOR=3.45, 95% CI: 1.56–7.63, p=0.002): Respondents with tertiary education were 3.45 times more likely to have a positive attitude compared to those with no formal education.
- Higher income (AOR=2.89, 95% CI: 1.34–6.21, p=0.007): Those earning above ZMW 5,000 were nearly three times more likely to have a positive attitude compared to those earning below ZMW 2,000.
- Knowledge score (AOR=1.42, 95% CI: 1.12–1.79, p=0.003): For each one-point increase in knowledge score, the odds of having a positive attitude increased by 42%. Knowledge score was treated as a continuous variable in the regression model to assess the dose-response relationship between knowledge and attitudes.
- Plastered/painted walls (AOR=2.12, 95% CI: 1.08–4.17, p=0.029): Respondents living in houses with plastered/painted walls were twice as likely to have a positive attitude compared to those with mud brick walls.
- Past spraying experience (AOR=4.56, 95% CI: 2.12–9.81, p<0.001): Those who had previously allowed their houses to be sprayed were 4.56 times more likely to have a positive attitude.
- Trust in health workers (AOR=1.38, 95% CI: 1.05–1.82, p=0.021): For each onepoint increase in trust in health workers, the odds of having a positive attitude increased by 38%. Trust in health workers was measured using a single Likert scale item: ‘I trust health workers who conduct IRS’. This was treated as a continuous variable (1=strongly disagree to 5=strongly agree) in the regression model

### Model Fit

- Hosmer-Lemeshow Test: ² = 6.34, p = 0.609 (model fit adequate)
- Nagelkerke R² = 0.342 (model explains 34.2% of variance)
- Overall Model Prediction Accuracy: 78.4%

### Summary of Key Findings

**Table 11.**
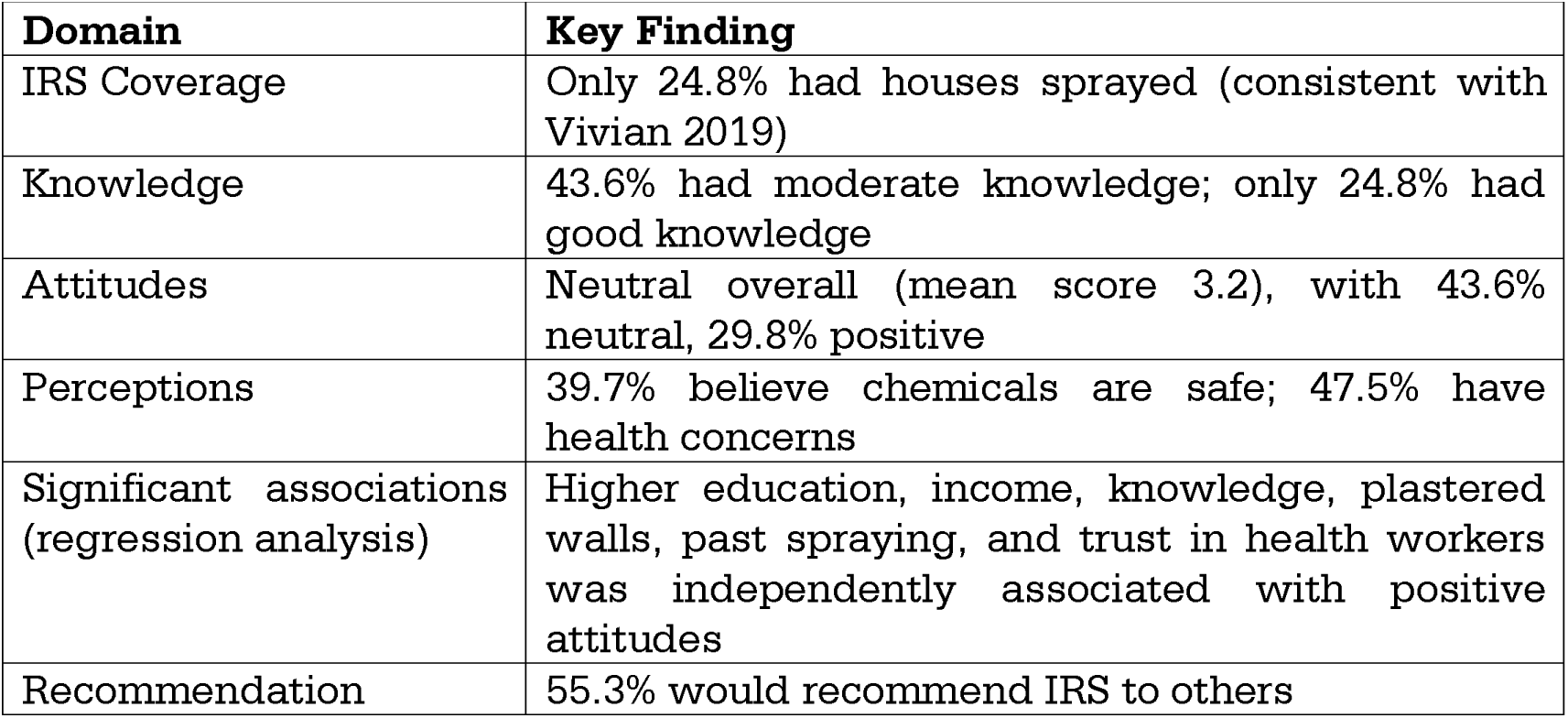
Summary of Key Findings.

| Domain | Key Finding |
| --- | --- |
| IRS Coverage | Only 24.8% had houses sprayed (consistent with Vivian 2019) |
| Knowledge | 43.6% had moderate knowledge; only 24.8% had good knowledge |
| Attitudes | Neutral overall (mean score 3.2), with 43.6% neutral, 29.8% positive |
| Perceptions | 39.7% believe chemicals are safe; 47.5% have health concerns |
| Significant associations (regression analysis) | Higher education, income, knowledge, plastered walls, past spraying, and trust in health workers was independently associated with positive attitudes |
| Recommendation | 55.3% would recommend IRS to others |

## DISCUSSION

### Principal Findings

This study reveals that IRS coverage in Masala (24.8%) is substantially below the WHO-recommended 80% threshold. Only 24.8% of respondents demonstrated good knowledge, and just 39.7% believed IRS chemicals are safe for humans. Fear of side effects (46.2%) and lack of trust in the spraying process (31.6%) were the most commonly reported barriers to acceptance. Higher education, higher income, better knowledge, plastered walls, past spraying experience, and trust in health workers were independently associated with positive attitudes towards IRS. However, given the cross-sectional design, these associations should not be interpreted as causal.

### Comparison with Other Studies

The IRS coverage of 24.8% observed in this study is nearly identical to the 24.8% reported by Vivian (2019) in the same community, suggesting that coverage has remained stagnant over the past several years. This coverage is substantially lower than the WHO-recommended 80% threshold (World Health Organization, 2021) and significantly below rates reported in other Zambian settings. Aongola et al. (2022) found higher acceptance rates in Luangwa district, where 67.9% of households accepted IRS. Similarly, Mkosha et al. (2024) reported that IRS coverage in Luwingu District was 64.4%. This disparity may reflect differences in program implementation—Luangwa and Luwingu are rural districts with targeted IRS programs, while Masala’s peri-urban setting may face unique challenges including population mobility, diverse housing types, and competing health priorities that complicate IRS delivery.

The knowledge levels found in this study (43.6% moderate, 24.8% good) are comparable to findings from other studies. Ngoma et al. (2022) similarly found that while general awareness of malaria prevention was high in peri-urban Ndola, detailed knowledge of specific interventions like IRS was limited. Jumbam et al. (2020) found moderate knowledge levels regarding malaria interventions in rural Zambia, with health workers and radio being primary information sources. Musachi (2017) similarly found that health workers were the most trusted source of information on malaria prevention in Solwezi.

The finding that fear of side effects was the most common reason for refusal (46.2%) aligns with studies from other settings. Musoke et al. (2015) reported that concerns about chemical exposure and health risks were major barriers to IRS acceptance in Uganda. Buttenheim et al. (2013) documented similar concerns in Peru, where mistrust of spraying chemicals was associated with lower participation in vector control campaigns. The perception that IRS causes unpleasant odors (mean=3.8) and leaves stains on walls (mean=3.6) found in this study is consistent with findings from Rodriguez et al. (2006) who documented similar aesthetic concerns in Mexico.

### Knowledge-Behavior Gap

A notable finding of this study is the knowledge-behavior gap: while 70.2% had heard about IRS and 55.3% believed it is effective, only 24.8% had ever allowed spraying in the last 12 months. This gap is consistent with Ngoma et al. (2022), who documented a similar pattern in peri-urban Ndola, where high awareness of malaria prevention did not translate into consistent practice.

Several factors may help explain this gap. First, knowledge about IRS may be superficial—knowing that IRS is effective does not guarantee understanding of how it works, what to expect during spraying, or how to prepare for the process. Second, perceived barriers (fear of side effects, health concerns, aesthetic issues) may outweigh knowledge in decision-making, particularly when individuals weigh potential benefits against perceived risks. Third, household decisionmaking dynamics may mean that individual knowledge does not translate into household acceptance if other family members object to the intervention.

This gap suggests that knowledge alone may be insufficient to significantly affect behavior change, and that structural and perceptual barriers likely play important roles. This mirrors findings from other health domains in Zambia. For instance, Mapiki et al. (2023) found that knowledge of hypertension management was insufficient to significantly affect adherence without practical, contextualized support—a pattern that appears to hold true for malaria prevention as well. This suggests that health education should evolve from information dissemination to skills-based, practical counseling that addresses real-world constraints, including safety reassurance, managing aesthetic concerns, and navigating household structural challenges (Matilda et al 2026).

### Perceptions of Chemical Safety

The finding that only 39.7% of respondents believed IRS chemicals are safe for humans is concerning. This is consistent with a growing body of evidence linking safety perceptions to intervention acceptance. Ocan et al. (2025), in their systematic review and meta-analysis, identified safety concerns as the most frequently cited reason for IRS non-acceptance across sub-Saharan Africa. The high proportion of respondents expressing health concerns (47.5%) and environmental concerns (31.6%) suggests that risk communication strategies may need to be strengthened.

The mechanisms behind these perceptions may vary by setting. In Masala, the high proportion of respondents citing health concerns suggests that risk communication during sensitization campaigns may be insufficiently addressing community anxieties. Previous research has shown that simply providing information may be less effective than involving community leaders and trusted health workers in two-way communication that acknowledges and addresses concerns (Atkinson et al., 2010). The effectiveness of sensitization campaigns may also be constrained by their timing and reach. In Masala, 29.8% of respondents had never heard about IRS, suggesting that sensitization campaigns may not be reaching all households. Furthermore, the finding that only 39.7% believed chemicals are safe suggests that those who are reached may not be receiving accurate risk communication.

The Zambian National Malaria Control Programme has emphasized community sensitization before each spray cycle (Ministry of Health, 2021), but this study suggests that existing sensitization efforts may not be effectively addressing safety concerns. Programs should consider pre-spraying household visits, visual aids demonstrating safety, engagement of local leaders, and direct dialogue with communities to address safety concerns comprehensively.

### Factors Associated with Positive Attitudes

#### Education and Income

The association between education and positive attitudes (AOR=3.45, p=0.002) is consistent with studies demonstrating that education is associated with improved health literacy and acceptance of public health interventions. The association between higher education and greater IRS acceptance aligns with findings from Aongola et al. (2022) in Luangwa and Jumbam et al. (2020) in rural Zambia. The association between higher income and positive attitudes (AOR=2.89, p=0.007) likely reflects the intersection of socioeconomic status with health literacy, access to information, and perhaps greater capacity to address practical barriers such as preparing homes for spraying.

This study’s findings on the importance of education, income, and trust in health workers align with broader reproductive health research in Ndola. Mbewe et al. (2026) found that among injectable contraceptive users at a peri-urban clinic, similar sociodemographic factors—including educational attainment and household income—were significantly associated with continued use. Their study also highlighted that provider-client communication and trust in health workers were critical for contraceptive continuity, reinforcing our observation that trust is a key associated factor of health intervention acceptance across different health domains in this setting.

#### Knowledge

The dose-response relationship between knowledge and attitudes (AOR=1.42 per one-point increase, p=0.003) underscores the importance of health education as a foundation for acceptance. However, the modest effect size suggests that knowledge alone may be insufficient—consistent with the knowledge-behavior gap discussed earlier. This implies that interventions should combine information provision with practical reassurance and community engagement.

#### Past Spraying Experience

Past spraying experience was strongly associated with positive attitudes (AOR=4.56, p<0.001). This finding is consistent with Buttenheim et al. (2013), who demonstrated that participation in vector control campaigns was associated with increased willingness to participate in future campaigns. This highlights the importance of achieving high initial coverage to create positive feedback loops. Conversely, it also suggests that communities with low coverage may remain in a cycle of non-participation, where lack of experience reinforces negative perceptions. Breaking this cycle may require targeted efforts to encourage first-time participation through intensive sensitization and reassurance.

#### Wall Material

The association between wall material and attitudes (AOR=2.12, p=0.029) may reflect both physical and social factors. Structurally, plastered walls retain insecticide more visibly and may provide a more obvious cue of protection, while mud brick walls absorb insecticide more readily, potentially reducing visible evidence of spraying and perceived effectiveness (Chebude et al., 2020). Socially, plastered walls are often associated with higher socioeconomic status, which may correlate with greater health literacy and trust in health interventions (Solomon et al., 2019). Programmatically, this suggests that households with mud brick walls may require additional reassurance and tailored messaging to address their specific concerns.

#### Trust in Health Workers

Trust in health workers was associated with positive attitudes (AOR=1.38, p=0.021), underscoring the importance of community engagement. This aligns with studies that have identified trust in health workers as a key factor associated with health intervention acceptance (Amin et al., 2021). In Zambia, Chanda et al. (2008) emphasized the importance of community sensitization and trust-building for successful IRS implementation. This suggests that health workers should be at the center of sensitization efforts. Training health workers in effective communication, addressing community concerns, and involving them in community engagement activities could strengthen trust and increase acceptance.

### Implications for Policy and Practice

These findings have several implications for policy and practice in Zambia.

First, IRS coverage in Masala (24.8%) is substantially below the WHO-recommended 80% threshold (World Health Organization, 2021), and targeted efforts are needed to increase coverage. The Ministry of Health and the National Malaria Elimination Centre should prioritize Masala for enhanced IRS outreach, including:

i. Pre-spraying household visits by community health workers to demonstrate safety and address concerns
ii. Involvement of community leaders as “IRS champions” to model acceptance
iii. Targeted messaging for households with mud brick walls and lower education levels
iv. A feedback mechanism for households to report concerns about the spraying process

Second, trust-building is important. The association between trust in health workers and positive attitudes suggests that health workers should be at the center of sensitization efforts. Training health workers in effective communication, addressing community concerns, and involving them in community engagement activities could strengthen trust and increase acceptance (Atkinson et al., 2010).

Third, structural barriers should be addressed. The association between wall material and attitudes suggests that households with mud brick walls may need tailored messaging. This could include reassuring households about safety, explaining that insecticide is safe for all wall types, and addressing aesthetic concerns. The concern about stains and odors (mean=3.8 and 3.6 respectively) suggests that communicating about these aspects before spraying could reduce resistance.

Fourth, the knowledge-behavior gap suggests that health education should go beyond information provision. As Mapiki et al. (2023) demonstrated, practical, contextualized support may be needed to shift behavior change. This could include demonstrating safety, addressing myths and misconceptions, and providing reassurance about side effects.

Implementation of these recommendations would require investment in community health worker training, sensitization materials, and monitoring. However, these costs should be weighed against the costs of maintaining low IRS coverage, which include continued malaria transmission, healthcare costs, and lost productivity. Of these recommendations, improving safety communication and building trust in health workers should be prioritized first, as these address the most commonly cited barriers and can be implemented with relatively modest additional resources.

### Strengths and Limitations

This study has several strengths. The use of a validated questionnaire, adequate sample size (n=282), systematic random sampling to minimize selection bias, high response rate (89.5%), and standardized data collection using KoBoToolbox enhance the validity and reliability of findings. The focus on a peri-urban community that is understudied yet representative of urban malaria transmission dynamics in Zambia provides valuable evidence for program planning.

Limitations should be acknowledged. First, the cross-sectional design precludes establishing causal relationships; we can only identify associations. Second, reliance on self-reported data may be subject to social desirability bias, potentially leading to over-reporting of positive attitudes. Enumerators were trained to build rapport and assure participants of confidentiality to minimize this effect. Third, the study was confined to one community, limiting generalizability to other settings. Fourth, recall bias may have affected responses regarding past spraying experience. Fifth, the use of a knowledge score based on self-reported awareness may not fully capture nuanced understanding of IRS. Sixth, the study did not collect data on actual malaria prevalence, which would have allowed analysis of associations between knowledge, attitudes, and health outcomes.

Seventh, the cross-sectional design captures a single point in time, precluding assessment of how attitudes evolve over time. Eighth, seasonal variation in malaria burden may influence perceptions, and data collection was limited to a single time period. Ninth, the study did not include qualitative methods to explore the depth of community perceptions. Tenth, sampling was limited to households with permanent structures, potentially excluding more transient populations who may have different perceptions and experiences with IRS.

Lastly, the inclusion of past spraying experience as a predictor of positive attitudes may be subject to reverse causality. While our cross-sectional design precludes determining temporal order, it is plausible that positive attitudes lead to participation, just as participation may reinforce positive attitudes. We therefore interpret this association as correlational rather than causal.

### Unanswered Questions and Future Research

Several important questions warrant further investigation. First, what specific misconceptions about IRS safety persist (e.g., does “health risk” mean acute illness, cancer, or something else)? Second, how do intra-household decisionmaking dynamics influence IRS acceptance (e.g., are male partners more likely to oppose spraying)? Third, what are the most effective communication channels and formats for addressing safety concerns (e.g., visual aids, demonstrations, testimonials)? Fourth, how do community health worker-based sensitization programs compare with mass media campaigns in changing attitudes and behavior?

Methodologically, future studies should employ mixed methods, combining quantitative surveys with qualitative interviews to capture the depth of community perceptions. Longitudinal studies are needed to track how knowledge and attitudes evolve with continued IRS campaigns. Implementation research is also needed to evaluate the effectiveness of specific interventions (e.g., pre-spraying household visits, community leader engagement) in improving coverage. Additionally, studies should assess the cost-effectiveness of different sensitization approaches to guide resource allocation. Finally, research is needed to understand how to address the needs of households with mud brick walls and other structural barriers to IRS acceptance.

### Conclusion

IRS coverage in Masala is low at 24.8%, substantially below the WHO-recommended 80% coverage threshold. Knowledge levels are moderate, with only 24.8% demonstrating good knowledge. Attitudes are neutral overall, and perceptions of chemical safety are poor, with only 39.7% believing IRS chemicals are safe for humans. Fear of side effects (46.2%) and lack of trust in the spraying process (31.6%) were the most commonly reported barriers to acceptance. Higher education, higher income, better knowledge, plastered walls, past spraying experience, and trust in health workers were associated with positive attitudes.

These findings have implications for policy and practice. First, targeted efforts are needed to increase IRS coverage through door-to-door sensitization, addressing safety concerns, and engaging community leaders. Second, trust-building is important, with health workers at the center of sensitization efforts. Third, structural barriers should be addressed through tailored messaging for households with different wall types. Fourth, health education should evolve from information dissemination to skills-based, practical counseling. Without such comprehensive approaches, malaria transmission in Masala is likely to persist, undermining Zambia’s goal of malaria elimination by 2026.

## WHAT IS ALREADY KNOWN ON THIS TOPIC

i. Indoor Residual Spraying is a proven vector control method that can reduce malaria transmission by up to 62% when implemented effectively (3).
ii. Acceptance of IRS varies across communities, influenced by knowledge, cultural beliefs, safety concerns, and socioeconomic factors (12,13).
iii. In Zambia, IRS has been implemented since the early 2000s as part of the Integrated Vector Management strategy, but coverage varies across districts (8).
iv. A study in Masala, Ndola found that only 24.8% of residents had their houses sprayed in the past 12 months (15).

## WHAT THIS STUDY ADDS

i. This study provides up-to-date evidence on the knowledge, attitudes, and perceptions towards IRS in Masala, confirming that coverage remains low at 24.8%.
ii. Fear of side effects (46.2%) and lack of trust in the spraying process (31.6%) are the primary barriers to acceptance, with safety concerns widespread (only 39.7% believe chemicals are safe).
iii. Higher education, higher income, better knowledge, plastered walls, past spraying experience, and trust in health workers was independently associated with positive attitudes towards IRS.
iv. Targeted community sensitization addressing safety concerns, strengthening trust in health workers, and providing practical reassurance are urgently needed to improve IRS uptake in urban Zambian communities.

## Data Availability

All data produced in the present study are available upon reasonable request to the authors

## REFERENCES

Amin ME, Akodu SO, Uzoechi N. Trust in health workers and acceptance of public health interventions. BMC Public Health. 2021;21(1):1–10.

Aongola M, Kaonga P, Michelo C, Zgambo J, Lupenga J, Jacobs C. Acceptability and associated factors of indoor residual spraying for malaria control by households in Luangwa district of Zambia: A multilevel analysis. PLOS Glob Public Health. 2022;2(8):e0000368.

Bhatt S, Weiss DJ, Cameron E, Bisanzio D, Mappin B, Dalrymple U, et al. The effect of malaria control on Plasmodium falciparum in Africa between 2000 and 2015. Nature. 2015;526(7572):207–211.

Buttenheim A, Paz Soldan V, Barbu C, Skovira C, Calderón J, Riveros L, et al. Is participation contagious? Evidence from a household vector control campaign in urban Peru. J Epidemiol Community Health. 2013;68(2):103–109.

Chanda E, Masaninga F, Coleman M, Sikaala C, Katebe C, Macdonald M, Baboo KS, Govere J, Manga L. Integrated vector management: the Zambian experience. Malar J. 2008 Aug 27;7:164. doi: 10.1186/1475-2875-7-164. PMID: 18752658; PMCID: PMC2551620.

Chanda, E., Chanda, J., Kandyata, A., Phiri, F.N., Muzia, L., Haque, U. and Baboo, K.S., 2013. Efficacy of ACTELLIC 300 CS, pirimiphos methyl, for indoor residual spraying in areas of high vector resistance to pyrethroids and carbamates in Zambia. Journal of medical entomology, 50(6), pp.1275–1281.

Grant M, Wilford A, Haskins L, Phakathi S, Mntambo N, Horwood CM. Trust of community health workers influences the acceptance of community-based maternal and child health services. Afr J Prim Health Care Fam Med. 2017 May 29;9(1):e1–e8. doi: 10.4102/phcfm.v9i1.1281. PMID: 28582988; PMCID: PMC5458568.

Jumbam DT, Stevenson JC, Matoba J, Grieco JP, Ahern LN, Hamainza B, et al. Knowledge, attitudes and practices assessment of malaria interventions in rural Zambia. BMC Public Health. 2020;20(1):216.

Mapiki C, Chalwe V, Chimfwembe K. Assessment of factors affecting adherence to antihypertensives in HIV clients at Chanyanya Rural Health Centre, Kafue, Zambia. J Sci Healthc Explor. 2023;8(1):1–12.

Mapiki, C.; Mulenga, M.; Mabaso, F.; et al. ‘Assessing the Knowledge, Attitudes and Perceptions on the Use of Intra-uterine Contraceptive Device Among Women in Ndola: A Case of Masala Clinic’. PREPRINT (Version 1) available at Research Square. 2026.

Matilda Mapiki, Chileleko Mapiki, Anna Phiri et al. Empowering Rural Learners: Assessing the Impact of Home Economics Education on Life Skills Acquisition in a Zambian Secondary School, 27 August 2026, PREPRINT (Version 1) available at Research Square [10.21203/rs.3.rs-10827405/v1]

Mbewe, A., Mapiki, C., & Mabaso, F. (2026). Beyond Uptake: High Discontinuation and Missed Reinjection Rates Among Injectable Contraceptive Users in Peri-Urban Ndola, Zambia. Preprints. 10.20944/preprints202608.1619.v1

Ministry of Health, Zambia. Copperbelt Malaria Elimination Program. Lusaka: Ministry of Health; 2021.

Mkosha M, Ngenda B, Nawa M. Factors associated with acceptance of indoor residual spraying among residents of Luwingu District, Northern Province of Zambia. Malar J. 2024;23(1):1–10.

Moss, W.J., Norris, D.E., Mharakurwa, S., Scott, A., Mulenga, M., Mason, P.R., Chipeta, J. and Thuma, P.E., 2012. Challenges and prospects for malaria elimination in the Southern Africa region. Acta tropica, 121(3), pp.207–211.

Musachi E. Preferences and perceptions about malaria prevention methods in Kimasala – Solwezi: A cross sectional study. Asian Pac J Health Sci. 2017;4(3):157–165.

Musoke D, Karani G, Ssempebwa JC, Etajak S, Guwatudde D, Musoke MB. Knowledge and practices on malaria prevention in two rural communities in Wakiso district, Uganda. Afr Health Sci. 2015;15(2):401–412.

Ngoma J, Haloka J, Chipoya G. Malaria knowledge and preventive practices in peri-urban households of Ndola, Zambia: A cross-sectional analysis of the knowledge-action gap. Int J Res Innov Soc Sci. 2026;10(6):12647–12655.

Ocan M, Ojiambo KO, Nakalembe L, Kinalwa G, Kinengyere AA, Nsobya S, et al. The effectiveness of indoor residual spraying for malaria control in sub-Saharan Africa: A systematic protocol review and meta-analysis. Int J Environ Res Public Health. 2025;22(6):822.

Oladipo HJ, Tajudeen YA, Oladunjoye IO, Yusuff SI, Yusuf RO, Oluwaseyi EM, et al. Increasing challenges of malaria control in sub-Saharan Africa: Priorities for public health research and policymakers. Sci Afr. 2022;81:104366.

Pluess B, Tanser FC, Lengeler C, Sharp BL. Indoor residual spraying for preventing malaria. Cochrane Database Syst Rev. 2010;(4):CD006657.

Rodríguez, Américo & Penilla-Navarro, R. & Rodríguez, Mario & Hemingway, Janet & Trejo Acevedo, Antonio & Hernandez-Avila, Juan Eugenio. (2006). Acceptability and perceived side effects of insecticide indoor residual spraying under different resistance management strategies. Salud Pública de México. 48. 317–24. 10.1590/S0036-36342006000400006.

Solomon, T., Loha, E., Deressa, W., Gari, T., Overgaard, H.J. and Lindtjørn, B., 2019. Low use of long-lasting insecticidal nets for malaria prevention in south-central Ethiopia: a community-based cohort study. PloS one, 14(1), p.e0210578.

Suuron, V.M., Mwanri, L., Tsourtos, G. and Owusu-Addo, E., 2020. An exploratory study of the acceptability of indoor residual spraying for malaria control in upper western Ghana. BMC public health, 20(1), p.465.

Vivian C (2019) Efficacy of Indoor Residual Household Spraying in Control of Malaria among Children Under Five Ndola, Zambia Texila International Journal of Public Healthhttps://www.academia.edu/41699666/Efficacy_of_Indoor_Residual_Household_Spraying_in_Control_of_Malaria_among_Children_Under_Five_Ndola_Zambia?sm=b&rhid=42430981290

World Health Organization. World malaria report 2021. Geneva: WHO; 2021.

World Health Organization. World malaria report 2023. Geneva: WHO; 2023.

Zambia Ministry of Health. Zambia National Malaria Indicator Survey 2022. Lusaka: Ministry of Health; 2023. https://www.nmec.org.zm/publications

Zambia National Malaria Elimination Centre. National Malaria Elimination Strategic Plan 2022-2024. Lusaka: Ministry of Health; 2022.

